# Comparison of Prehospital Vascular Access Strategies and Their Impact on Survival in Out-of-Hospital Cardiac Arrest

**DOI:** 10.1101/2025.06.19.25329959

**Authors:** Sheng-Min Lin, Cheng-Yu Chien, Chip-Jin Ng, Liang-Tien Chien, Hsin-Tzu Yeh, Pang-Ting Hsu, Ming-Fang Wang, Hsiao-Jung Tseng, Chien-Hsiung Huang

**Author notes:** Sheng-Min Lin and Cheng-Yu Chien contributed equally to this article. Correspondence to: Chien-Hsiung Huang, MD, PhD, Department of Emergency Medicine, Chang Gung Memorial Hospital, Linkou and College of Medicine, Chang Gung University, Taoyuan 333, No. 5 Fushing St., Gueishan Dist., Taoyuan City, Taiwan. # 2505.

## Abstract

**Background:** Out-of-hospital cardiac arrest (OHCA) remains a critical emergency with low survival rates despite advanced prehospital interventions. Early epinephrine administration, particularly in non-shockable rhythms, improves outcomes. While intravenous (IV) access is the standard route for drug delivery, it is often difficult to obtain in the prehospital setting. Intraosseous (IO) access offers a viable alternative, but its comparative survival benefit remains unclear. Few studies have examined the impact of IO access on outcomes relative to patients who received no prehospital vascular access. This study aims to assess survival outcomes among OHCA patients receiving different prehospital vascular access strategies.

**Methods and Results:** This retrospective cohort study included adult patients with non-traumatic OHCA in Taoyuan, Taiwan (June 2021–June 2024). Patients were categorized into four groups: IV, IO (humerus), failed IV, and no-access attempt. Primary outcomes were survival to discharge and favorable neurological status (CPC 1–2); secondary outcomes included prehospital ROSC and survival over 2 hours. Multivariable logistic regression adjusted for confounders. Among 4,424 patients, the IO group had the highest rates of ROSC (13.0%), 2-hour survival (28.0%), and discharge survival (16.6%), while the failed IV group had the lowest. IO access was associated with better outcomes than no-access attempt, with or without epinephrine. IO patients receiving epinephrine had the highest estimated survival and neurological outcomes. Each minute delay in epinephrine administration reduced survival odds by 4%.

**Conclusions:** Prehospital vascular access—especially IO—was linked to improved OHCA survival and neurological outcomes. Prompt IO access should be considered when IV attempts fail.

**Clinical perspective:** *What Is New?:*

- This is the first study to comparing vascular access including IV, IO, IV fails and no access attempt before arriving hospital on non-traumatic OHCA patients.
- In pre-hospital settings, IO access should be promptly established when IV fails, which significantly improving OHCA patient survival and neurological outcomes.

*What Are the Clinical Implications?:*

- The study emphasizes the importance of establishing IO access while IV access failed on OHCA patients prior to hospital arrival in order to improve survival and neurological outcomes.

## Background

Introduction Out-of-hospital cardiac arrest (OHCA) remains a significant public health challenge, with global incidence rates exceeding 56 per 100,000 person-years among adults. Despite extensive efforts, survival rates to hospital discharge remain low, typically under 8%, with fewer than 3% of survivors experiencing favorable neurological outcomes. [1–4] Timely interventions, including early cardiopulmonary resuscitation (CPR), defibrillation, and rapid administration of epinephrine, are critical elements in improving OHCA outcomes. [5–8] Timely administration of epinephrine is a critical component of advanced prehospital resuscitation performed by Emergency Medical Services (EMS) personnel. Early epinephrine administration in non-shockable OHCA has been associated with improved rates of return of spontaneous circulation (ROSC), survival to hospital discharge, and survival at three months post-arrest. [7] Delays in epinephrine administration have been linked to a 4% decrease in survival odds per minute from EMS arrival to drug administration. [8]

Intravenous (IV) access is the standard route for medication delivery during resuscitation; however, prehospital IV placement frequently encounters practical challenges, such as patient condition, limited space, and the chaotic environment. The success rate of IV access in OHCA patients varies across studies, with a Korean study reporting a 73.5% success rate [9], while a U.S. study found a first-attempt success rate of only 49%. [10] When IV access proves difficult, intraosseous (IO) access is recommended as an alternative by current guidelines, offering high success rates and rapid placement. [9–11] In a Canada study, the first-attempt success rate of IO access in adult OHCA was 97% in humerus group and 98% in tibia group, respectively. [11] Although IO access is widely accepted in clinical practice, evidence on its comparative effectiveness to IV access regarding survival and neurological outcomes remains inconclusive. [12] A recent study reported no differences in sustained ROSC rates or favorable neurological outcomes (cerebral performance category, CPC 1–2) between IO and IV access, even when comparing different insertion sites (upper vs. lower limbs). [13] In contrast, a study from Oregon, USA, found that both tibial and humeral IO access were associated with lower odds of ROSC upon emergency department arrival compared to IV access. [14] Yang’s study further indicated no significant difference in survival to hospital discharge or neurological outcomes between IV and IO access in non-traumatic OHCA patients, though better outcomes were observed when vascular access was established in the upper extremities. [15] Furthermore, the impact of failed IV attempts and the subsequent management strategy in OHCA cases needs further investigation. A recent study from Korea compared patients who successfully received IV access via prehospital epinephrine with those in the no IV attempt group. The results showed a higher survival-to-hospital discharge rate in the no IV attempt group but with poorer neurological outcomes. [9] This finding was influenced by a longer EMT transport time in the IV attempt group. Additionally, no significant differences were observed in survival to hospital discharge or neurological outcomes between the IV failure group and the no IV attempt group. [9]

In current practice, IV access is typically attempted first unless the patient presents with apparent difficulty in IV cannulation, in which case EMTs proceed directly to IO access. In some cases, after multiple failed IV attempts, EMTs may opt to expedite hospital transport without establishing alternative vascular access.

The impact of vascular access establishment on survival outcomes in OHCA patients remains unclear. Limited studies have investigated how IO accessibility influences survival, particularly in comparison to patients who did not receive prehospital vascular access. Therefore, this study aims to assess survival and neurological outcomes in OHCA patients receiving any strategies of vascular access, providing insights for optimal prehospital management strategies.

## Methods

### Study Design and Setting

The data that support the findings of this study are available from the first author on reasonable request. This study was a retrospective cohort analysis conducted in Taoyuan City, Taiwan, from June 2021 to June 2024.

Data were extracted from the Taoyuan OHCA registry, a prehospital database that systematically records out-of-hospital cardiac arrest (OHCA) cases based on the Utstein reporting guidelines. The EMS system in Taoyuan includes 41 EMS stations and 13 designated first-aid hospitals, all of which adhere to the 2020 American Heart Association (AHA) Advanced Cardiovascular Life Support (ACLS) guidelines.

The study protocol was reviewed and approved by the Hospital Ethics Committee on Human Research of Taiwan’s Chang Gung Medical Foundation (permit number: 202400158B0). Given the retrospective nature of the study, the requirement for informed consent was waived.

### Study Population

The study population consisted of adult patients (≥18 years old) who experienced non-traumatic OHCA during the study period and received prehospital resuscitation attempts by EMS personnel. Patients were excluded if they had a pre-existing do-not-resuscitate (DNR) order, exhibited signs indicating death at the scene (e.g., rigor mortis, decapitation, decomposition), or if OHCA was caused by drowning, hanging, or intoxication. Additionally, cases with pregnancy, missing essential prehospital or hospital data were excluded, as well as those in which IO vascular access was established in the lower extremities.

### Treatment Groups

All eligible patients were classified into four groups based on their prehospital vascular access. The No-access attempt Group included patients who did not receive or try to set a peripheral line or epinephrine during EMT resuscitation until arrive ED. The IV Group consisted of patients who successfully received IV access, while the IO Group comprised those who received humerus IO access. The Failed IV Group included patients in whom IV access attempts were unsuccessful in the field, and peripheral access was not successfully established until hospital arrival. The selection of vascular access was determined by EMS personnel based on the clinical conditions at the scene. For analysis purposes, classification was based on the final attempted vascular access route, reflecting the last route attempted before arrive ED. The establishment of peripheral pathways can be performed by EMT-intermediates, but the administration of epinephrine can only be given through EMT-Paramedics, so the administration of epinephrine cannot be given 100% of the IV or IO group.

### Outcome Measures

The primary study outcomes were survival to hospital discharge and neurologically favorable survival, defined as a CPC score of 1 or 2 at discharge. The secondary outcomes included prehospital ROSC and survival for at least two hours post-arrest.

### Data Collection and Variables

Patient demographic and clinical data were obtained from EMS run sheets and hospital records, including patient characteristics, arrest-related factors, EMS performance metrics, and hospital outcomes. Patient characteristics included age, sex, hospital level and medical history. Arrest-related factors encompassed witnessed arrest, bystander CPR, use of public automated external defibrillators (AEDs), and initial cardiac rhythm. EMS performance metrics comprised response time (from emergency call to EMS arrival), duration of on-scene resuscitation (scene time interval), transport time, airway management strategy, vascular access type, and time to epinephrine administration. Hospital outcomes were assessed based on survival status and the Cerebral Performance Category (CPC) score at discharge.

The time to epinephrine administration was defined as the interval from the emergency call to the first administration of epinephrine. For patients in whom IV access attempts were unsuccessful, the recorded time corresponded to the point when EMS personnel abandoned IV attempts.

### Statistical analysis

Descriptive statistics were reported as mean ± SD or median with interquartile range (IQR) for continuous variables, as appropriate, and as counts with percentages for categorical variables. To assess differences across treatment groups, one-way ANOVA was used, with the Kruskal-Wallis test applied when the normality assumption was violated. The Chi-square test examined differences in proportions among groups.

Logistic regression analysis was used to understand the association between different treatments and the outcomes: pre-hospital ROSC, survival over hours, survival to discharge, and good CPC. Adjusted odds ratios were reported as the main results, comparing IO or IV or failed IV groups with the no-access attempt group in a multivariable logistic regression model; covariates were included if they were significant in univariate analysis and potential confunders. Estimated probabilities and their 95% confidence intervals (CIs) were derived using multivariable logistic regression models. Each outcome—pre-hospital ROSC, survival over 2 hours, survival to hospital discharge, and favorable neurological outcome—was modeled separately. Statistical analysis was conducted using SAS version 9.4, and statistical plots were generated with the R package “ggplot2.” All tests were two-sided, with a p-value less than 0.05 considered statistically significant.

## Result

Between June 2021 and June 2024, 13,051 OHCA cases occurred in Taoyuan City. Of these, 7,765 were excluded due to on-scene death declarations or family-signed Do-Not-Resuscitate orders, and 862 additional cases were excluded due to specific criteria (e.g., pregnancy, age <18 years, intoxication, incomplete data, lower limb IO access). The final cohort comprised 4,424 patients divided into four groups: no-access attempt (n=2,229), IO access (n=656), IV access (n=1,278), and failed IV (n=261) (Figure 1).

**Figure 1.**
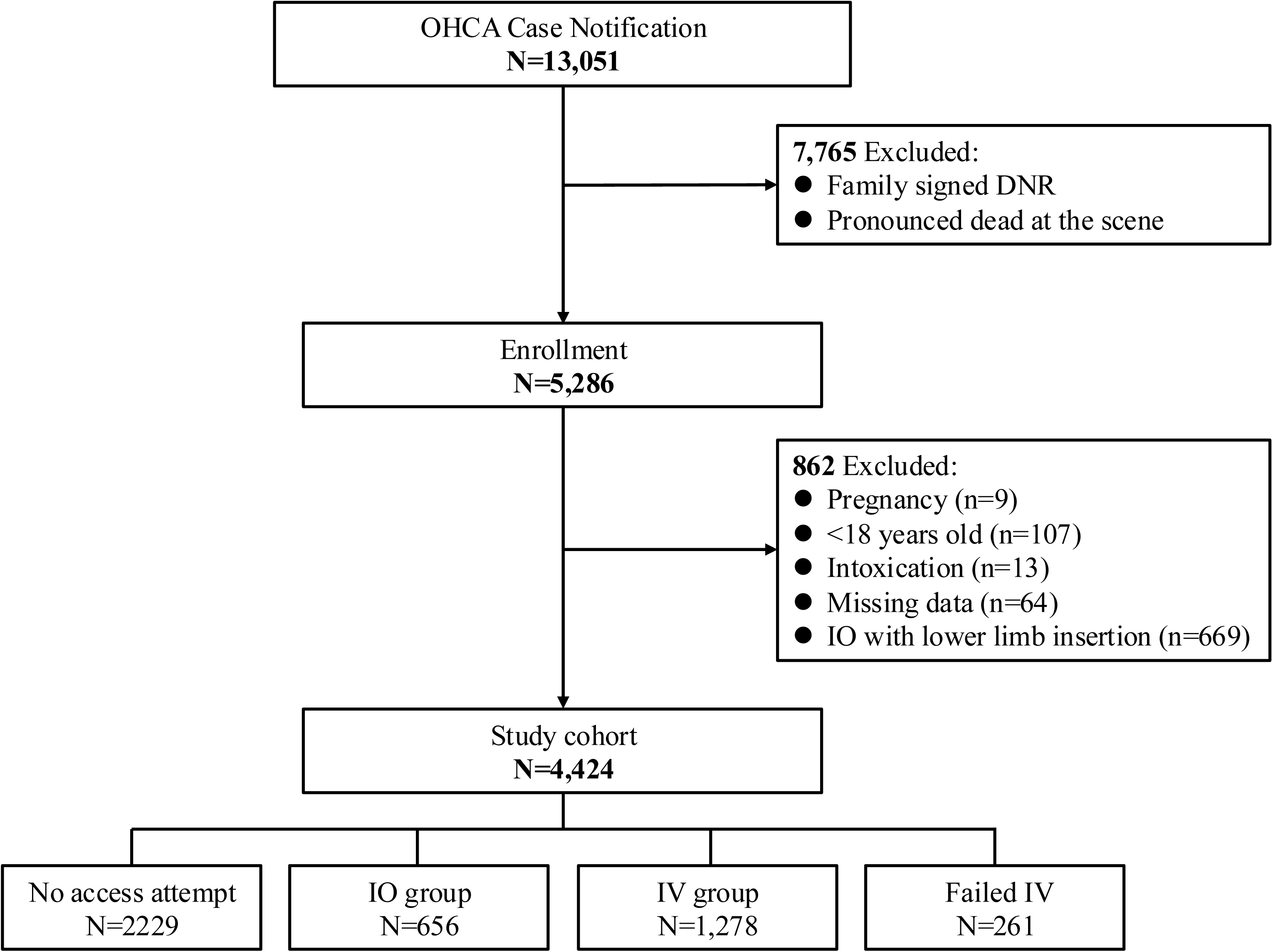
Flow chart of case selection process for the study

### Baseline characteristics of OHCA patients

Patients receiving IO were younger (mean 61.9 ± 17.15 years) than other groups. Witnessed arrests (41.7%) and bystander CPR (71.4%) were highest in the IV group. Shockable rhythms occurred more common in the IO (24.8%) and IV (21.5%) groups compared to the no-access attempt or failed-IV groups (p<0.001). The IO group had the shortest response time (median 5 min, IQR: 3–6 min) but longest scene time (median 17 min, IQR: 14–21 min; p<0.001). Epinephrine was administered in 66.0% (IO) and 58.2% (IV) groups, with longer time to epinephrine administration in IV (median 21 min) than IO (median 18 min) (Table 1).

**Table 1.**
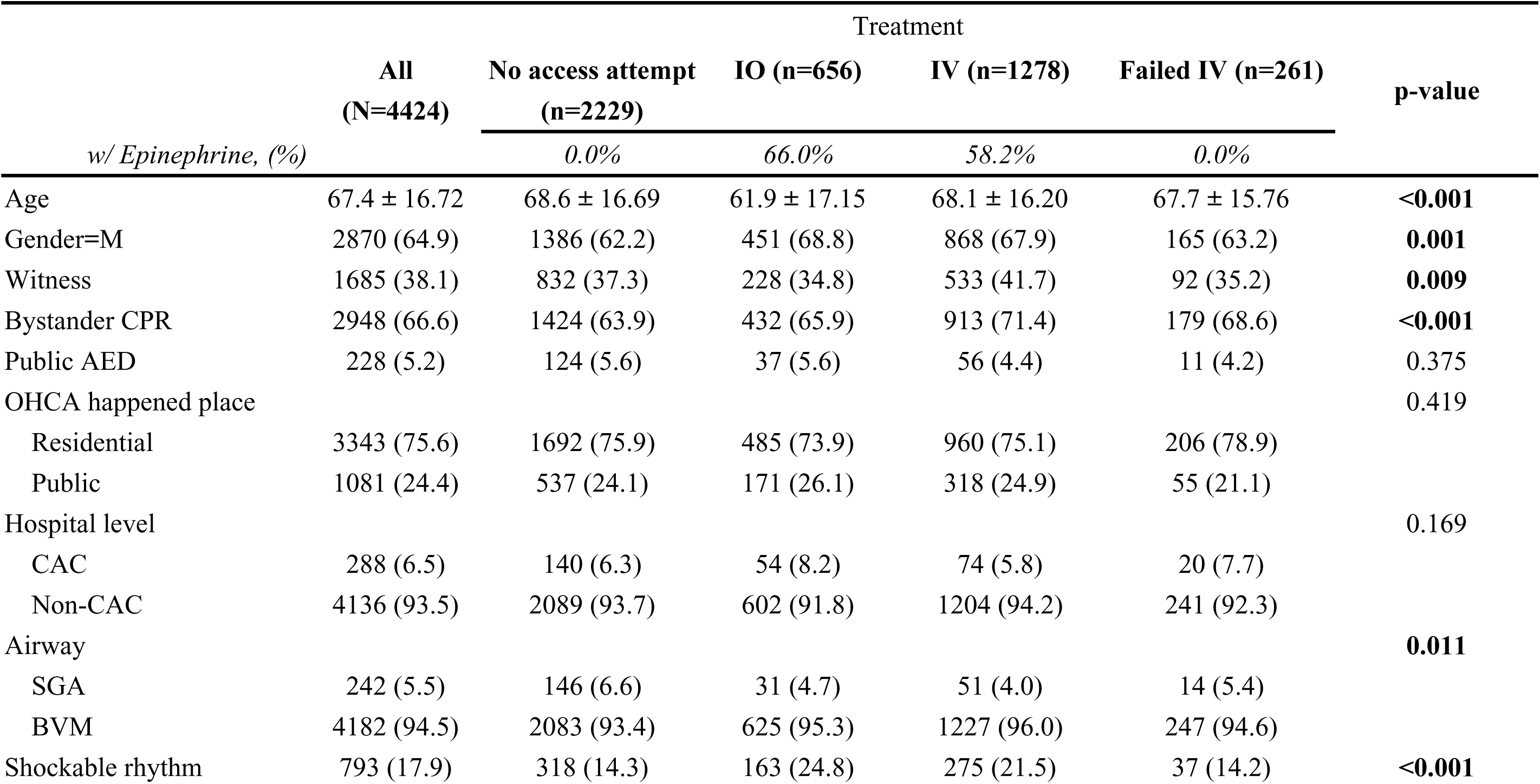
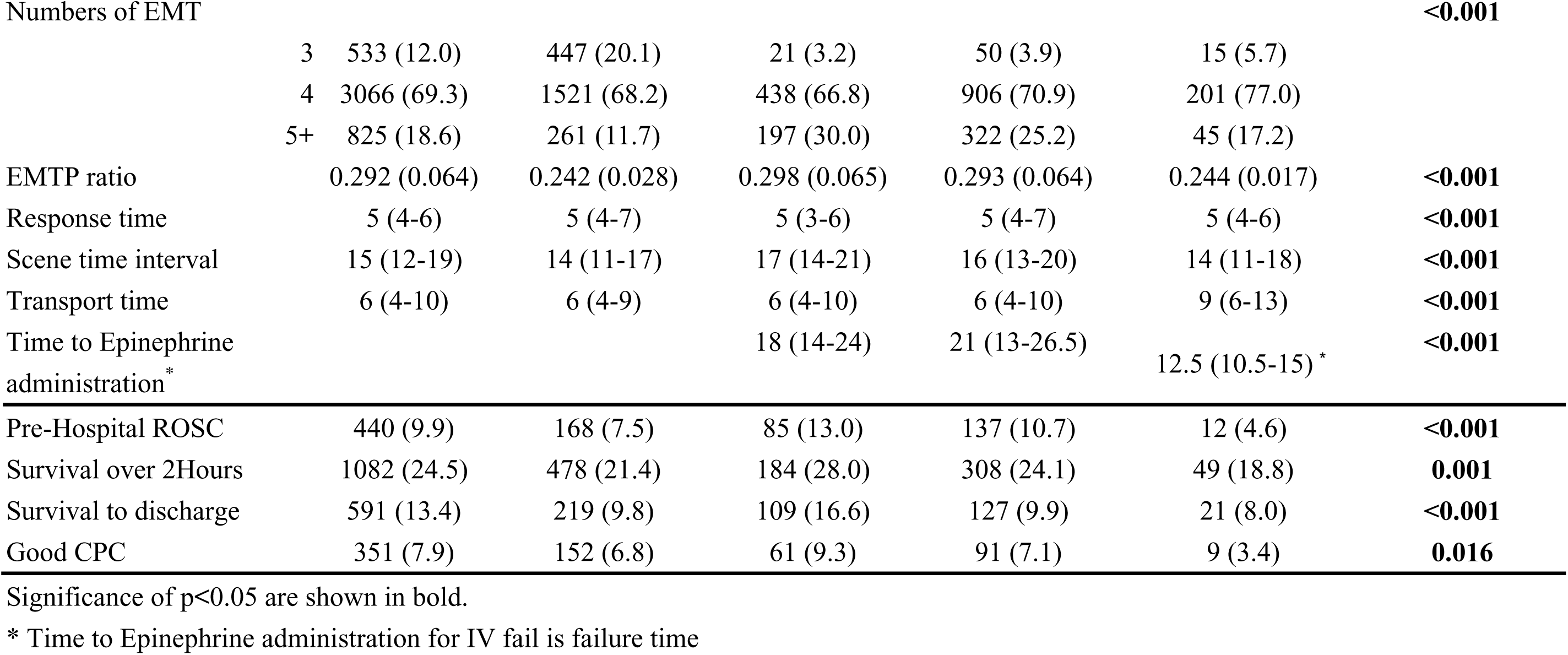
Descriptive statistics across different treatment groups.

### Outcomes assessment for different treatment groups

The IO group showed the highest rates of pre-hospital ROSC (13.0%), survival over 2 hours (28.0%), and survival to discharge (16.6%). The failed IV group had the lowest outcomes across all metrics (ROSC 4.6%, survival >2h 18.8%, discharge 8.0%, good CPC 3.4%) (Table 1).

Multivariate analysis (Table 2) revealed significantly higher odds in the IO group compared to no-access attempt group for ROSC (aOR 1.52, 95% CI: 1.07– 2.16), 2-hour survival (aOR 1.50, 95% CI: 1.17–1.93), and discharge (aOR 2.30, 95% CI: 1.69–3.12). The IV group had moderately higher odds for survival over 2 hours (aOR 1.22, 95% CI: 1.01–1.51, p=0.047) and survival to discharge (aOR 1.27, 95% CI: 1.03–1.67, p=0.042). Epinephrine use significantly increased odds of survival to discharge (aOR 1.67, 95% CI: 1.25–2.22, p<0.001) and good CPC (aOR 1.45, 95% CI: 1.11–2.04, p=0.041).

**Table 2.** Estimated Odds ratios for the Outcomes

| Comparison | Pre-hospital ROSC |  | Survival over 2 hours |  | Survival to discharge |  | Good CPC |  |
| --- | --- | --- | --- | --- | --- | --- | --- | --- |
|  | aOR(95% CI) | *p-value* | aOR(95% CI) | *p-value* | aOR(95% CI) | *p-value* | aOR(95% CI) | *p-value* |
| IO vs. no access attempt | 1.52(1.07-2.16) | **0.018** | 1.50(1.17-1.93) | **0.001** | 2.30(1.69-3.12) | **<0.001** | 1.27(0.87-1.87) | 0.221 |
| IV vs. no access attempt | 1.30(0.96-1.75) | 0.093 | 1.22(1.01-1.51) | **0.047** | 1.27(1.03-1.67) | **0.042** | 1.12(0.80-1.56) | 0.503 |
| Failed IV vs. no access attempt | 0.59(0.32-1.08) | 0.089 | 0.85(0.61-1.19) | 0.342 | 0.79(0.49-1.28) | 0.345 | 0.48(0.24-0.96) | **0.039** |
| w/ Epinephrine vs. w/o E. | 1.09(0.81-1.46) | 0.589 | 1.16(0.94-1.45) | 0.163 | 1.67(1.25-2.22) | **<0.001** | 1.45(1.11-2.04) | **0.041** |
| Time to Epinephrine administration | 0.95(0.93-0.98) | **<0.001** | 0.97(0.96-0.99) | **0.003** | 0.96(0.94-0.98) | **0.005** | 0.96(0.93-0.99) | **0.017** |

Significance of $p < 0.05$ are shown in bold.

### Stratification analysis

Stratified analysis by epinephrine use (Table 3) showed that with epinephrine, IO access was superior to IV, particularly in survival to discharge (aOR 0.52, IV vs IO) and good CPC (aOR 0.51, IV vs IO). Without epinephrine, IO demonstrated markedly improved outcomes over no-access attempt for all measures (ROSC, 2-hour survival, discharge, good CPC, all p<0.001). The failed IV group consistently showed poorer neurological outcomes. Estimated probabilities for outcomes were highest in IO patients receiving epinephrine (Figure 2, Supplementary Table S1-2), underscoring the effectiveness of IO access combined with timely epinephrine administration.

**Table 3.** Stratification analysis Comparison

| Comparison | pre-hospital ROSC |  | Survival over 2 Hours |  | Survival to discharge |  | Good CPC |  |
| --- | --- | --- | --- | --- | --- | --- | --- | --- |
|  | aOR (95% CI) | p-value | aOR (95% CI) | p-value | aOR (95% CI) | p-value | aOR (95% CI) | p-value |
| **w/ Epinephrine treatment** |  |  |  |  |  |  |  |  |
| IV vs IO | 0.86(0.58-1.28) | 0.465 | 0.75(0.56-1.01) | 0.055 | 0.52(0.33-0.82) | **0.005** | 0.51(0.29-0.89) | **0.019** |
| **w/o Epinephrine treatment** |  |  |  |  |  |  |  |  |
| IO vs. no access attempt | 2.08(1.39-3.12) | **<0.001** | 2.46(1.82-3.31) | **<0.001** | 4.66(3.36-6.47) | **<0.001** | 2.08(1.38-3.13) | **<0.001** |
| IV vs. no access attempt | 1.00(1.43-0.70) | 0.998 | 1.08(1.37-0.85) | 0.564 | 1.56(2.27-1.09) | **0.016** | 1.35(2.04-1.11) | **0.032** |
| Failed IV vs. no access attempt | 0.58(0.31-1.08) | 0.084 | 0.85(0.61-1.19) | 0.342 | 0.79(0.49-1.27) | 0.332 | 0.47(0.23-0.95) | **0.036** |

aOR, adjusted odds ratio.
Significance of $p < 0.05$ are shown in bold.

**Figure 2.**
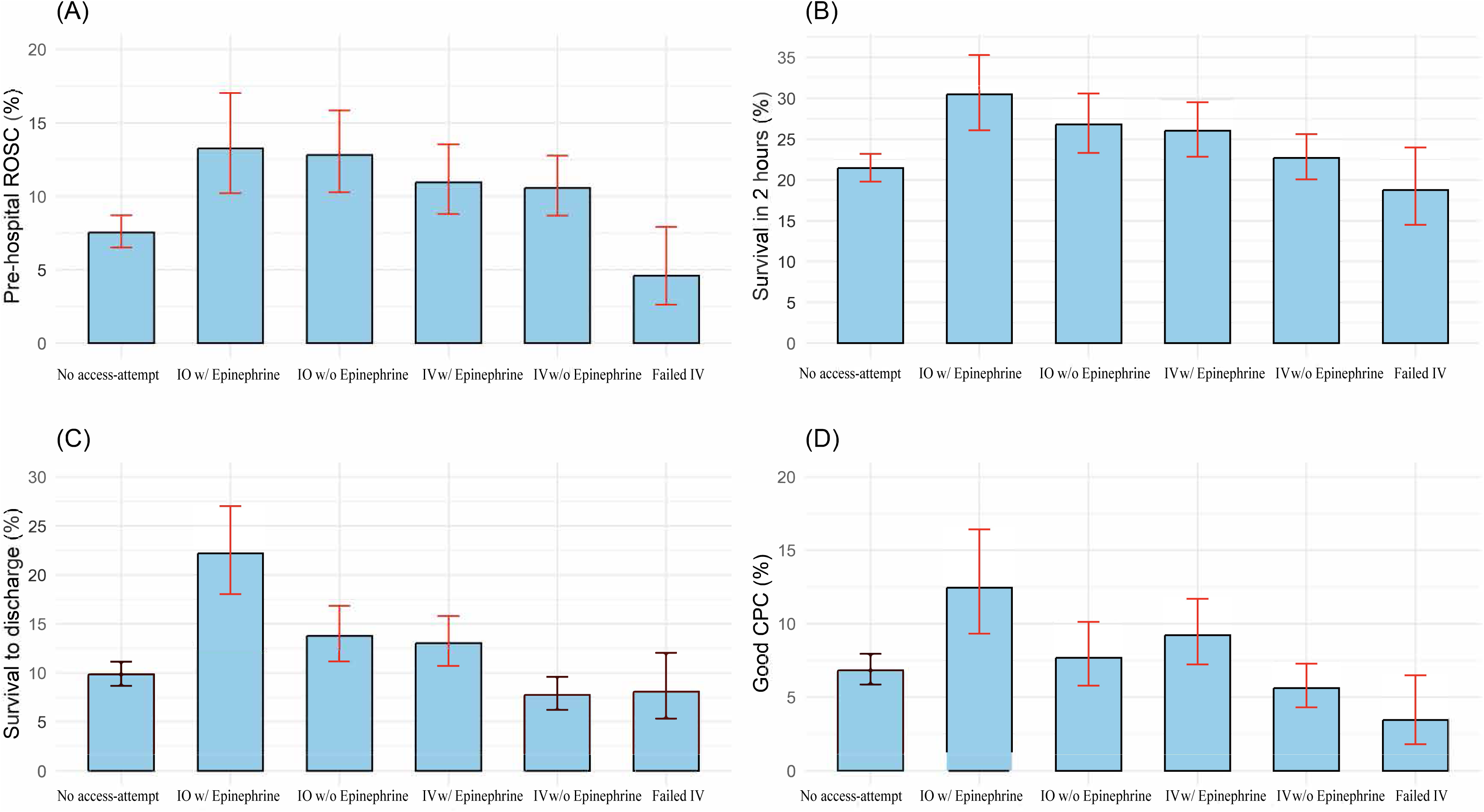
Estimated probabilities for (A)pre-hospital ROSC, (B) survival in (over) 2 hours, (C) survival to discharge and (D) good CPC.

## Discussion

Our findings indicate that in the prehospital cardiac arrest setting, when IV access attempts fail, IO access should be promptly established. Patients in the IO group demonstrated better survival outcomes than those in the no-access attempt group. In an additional analysis of prehospital epinephrine administration, OHCA patients exhibited improved survival and neurological outcomes (4% per minute). Moreover, the IO group had higher rates of survival to discharge, and good CPC compared to the IV group. Conversely, the IV failure group had worse outcomes than the no-access attempt group, with a significantly lower rate of good CPC. The survival prognosis of OHCA patients who did not receive any peripheral access and failed to establish one was generally poor, regardless of whether epinephrine was administered. In particular, OHCA patients who experienced failed IV attempts and received no further intervention had the worst prognosis. These findings underscore the critical role of establishing IO access when IV attempts fail in OHCA patients before hospital arrival.

According to our study, IO route establishing prior to hospital arrival stand as an important role than no-access attempt group, especially in the stratification analysis of non-epinephrine given group. IO route can substitute for peripheral line before hospital IV access is setting up. Even if epinephrine was not administered before hospital arrival, we still can give fluid challenge through IO access and medication was given via IO immediately after arrival at the hospital. [16] Our findings demonstrated significantly worse outcomes with IV epinephrine administration compared to IO delivery. This contrasts with three randomized controlled trials published in recent, which reported no significant difference in survival between IV and IO routes.[17–19] However, unlike those trials, there was no difference in the timing of IV OR IO epinephrine administration; our study observed a median 3-minute earlier administration of epinephrine via IO, supporting prior evidence that delays in epinephrine administration are associated with decreased survival rates. [15]

Timely administration of epinephrine is critical in cardiac arrest resuscitation. Current resuscitation guidelines recommend early epinephrine administration in patients with non-shockable rhythms to improve ROSC and neurological outcomes.[20] Several studies have demonstrated that epinephrine given within five minutes of collapse is associated with higher survival to hospital discharge in both shockable and non-shockable OHCA patients.[21] In traumatic OHCA, prehospital epinephrine use has also been linked to increased rates of sustained ROSC.[22] In our study, prehospital administration of epinephrine was associated with significantly higher rates of survival to discharge and favorable neurological outcomes compared to no epinephrine use. Prior research has reported a 4% decrease in survival for every one-minute delay in epinephrine administration.[23] Consistent with this, our findings showed that epinephrine was administered a median of three minutes earlier in the IO group compared to the IV group. In prehospital settings, IV access failure rates may be as high as 50%,[24] which could lead to delayed drug delivery. The high first-attempt success rate and rapid placement of IO access may reduce interruptions in chest compressions and delays in defibrillation analysis or the administration of medication, thereby contributing to a more efficient and uninterrupted resuscitation process.

The IV failure group had a poorer CPC rate than the treatment group. There was longer transporting time in the IV failure group than the other groups whose first post-hospital epinephrine administration may have been more delayed. Failure of intravenous catheter placement increased if visual identification of the target vein by palpating the extremity was impossible. [25] The situation is similar to OHCA patients whose vascular lumen might be collapsed due to poor perfusion, resulting in an increasing failure rate of IV access. Under the IV failure circumstances, the patient must be speculated to be more severe. Thus, IO access might be applicable despite the failure of the IV route, which might subsequently bridge in-hospital treatment. Our findings support protocol optimization emphasizing early IO access, particularly when IV attempts are likely to fail, to ensure timely drug delivery and minimize delays in resuscitation.

### Limitations

This study has several limitations. First, as a retrospective cohort study, it is inherently subject to bias and unmeasured confounders. To minimize this, we adjusted for multiple variables in the multivariable logistic regression model and conducted stratified analyses based on epinephrine administration. Second, the choice of IV or IO access was determined by EMS personnel based on real-time clinical judgment, which may have influenced the observed outcomes. To address this, we categorized patients according to the final attempted vascular access route and analyzed baseline characteristics to ensure comparability between groups. Third, we excluded cases in which IO access was established in the lower extremities due to concerns regarding differences in drug pharmacokinetics and resuscitation efficacy. While this limits the generalizability of our findings to all IO access sites, prior studies have suggested superior outcomes with upper extremity IO access, supporting our approach. Fourth, data on CPR quality, including compression depth, rate, and hands-off time, were not available in our dataset. However, we included key EMS performance metrics, such as response time and on-scene time, as indirect indicators of CPR process efficiency. Finally, this study was conducted in a single urban EMS system in Taiwan, which may limit the generalizability of our findings to other regions. Nevertheless, the study included a large OHCA cohort with standardized data collection based on the Utstein guidelines, improving internal validity. Furthermore, our findings are consistent with prior research, reinforcing the importance of prompt IO access when IV attempts fail. Future studies with prospective designs and multi-center validation are warranted to confirm these findings across different EMS settings.

## Conclusions

Establishing either intraosseous or intravenous access before hospital arrival was associated with significantly better survival prognosis than providing no prehospital vascular access. IO access should be promptly established when IV fails, significantly improving OHCA patient survival and neurological outcomes. Avoiding unnecessary IV attempts and promptly transitioning to IO access may enhance overall resuscitation efficiency and improve survival and neurological outcomes in prehospital settings.

## Declaration of Competing Interest

None.

The authors of this review will not be submitting a manuscript on Comparison of Prehospital Vascular Access Strategies and Their Impact on Survival in Out-of-Hospital Cardiac Arrest to another journal until JAHA makes a decision to reject or actually publishes (not just accepts) this manuscript.

No AI program was utilized in the construction of this manuscript.

## Data Availability

The data that support the findings of this study are available from the first author on reasonable request.

## Acknowledgments

The authors thank the staff and participants of the study for their important contributions.

## Nonstandard Abbreviations and Acronyms

OHCA: Out-of-hospital cardiac arrest
IV: Intravenous
IO: Intraosseous
EMS: Emergency Medical Services
CPR: Cardiopulmonary resuscitation
ROSC: Return of spontaneous circulation
CPC: Cerebral performance category
DNR: Do-not-resuscitate
AEDs: Automated external defibrillators
IQR: Interquartile range
CIs: Confidence intervals

## Supplementary Tables

**Supplementary Table S1.** Univariate analysis: the estimated odds ratio by simple logistic regression.

|  | pre hospital ROSC |  | Survival over 2 hours |  | Survival to discharge |  | Good CPC |  |
| --- | --- | --- | --- | --- | --- | --- | --- | --- |
|  | OR(95% CI) | p-value | OR(95% CI) | p-value | OR(95% CI) | p-value | OR(95% CI) | p-value |
| Treatment group |  |  |  |  |  |  |  |  |
| No access attempt | Reference |  | Reference |  | Reference |  | Reference |  |
| IO | 1.83(1.38-2.41) | **<0.001** | 1.43(1.17-1.74) | **<0.001** | 1.83(1.43-2.34) | **<0.001** | 1.40(1.03-1.91) | **0.033** |
| IV | 1.47(1.16-1.87) | **0.001** | 1.16(1.04-1.37) | **0.043** | 1.34(1.01-1.67) | **0.032** | 1.05(0.80-1.37) | 0.735 |
| Failed IV | 0.59(0.32-1.86) | 0.086 | 0.85(0.61-1.17) | 0.318 | 0.80(0.50-1.28) | 0.358 | 0.49(0.25-0.97) | **0.040** |
| w/ Epinephrine vs. w/o E | 1.43(1.15-1.78) | **0.001** | 1.09(0.94-1.28) | 0.262 | 1.54(1.11-1.91) | **0.021** | 1.56(1.13-1.96) | **<0.001** |
| Age | 0.98(0.98-0.99) | **<0.001** | 0.99(0.98-0.99) | **<0.001** | 0.98(0.97-0.98) | **<0.001** | 0.97(0.96-0.98) | **<0.001** |
| Gender (F vs. M) | 1.00(0.80-1.24) | 0.982 | 0.99(0.86-1.15) | 0.944 | 0.98(0.80-1.19) | 0.823 | 0.82(0.64-1.05) | 0.112 |
| Witness (Y vs. N) | 3.42(2.76-4.24) | **<0.001** | 2.68(2.32-3.09) | **<0.001** | 3.22(2.64-3.92) | **<0.001** | 3.18(2.50-4.04) | **<0.001** |
| Public vs. Residential | 0.96(0.77-1.20) | 0.729 | 1.06(0.92-1.23) | 0.404 | 0.98(0.80-1.20) | 0.852 | 0.90(0.71-1.16) | 0.424 |
| CAC vs. non-CAC | 1.96(1.35-2.85) | **<0.001** | 1.26(0.94-1.71) | 0.126 | 1.90(1.33-2.70) | **<0.001** | 2.18(1.47-3.25) | **<0.001** |
| BVM vs. SGA | 1.93(2.39-1.56) | **<0.001** | 1.88(2.19-1.61) | **<0.001** | 2.42(2.95-1.99) | **<0.001** | 2.77(3.50-2.19) | **<0.001** |
| AED use (Y vs. N) | 0.51(0.36-0.71) | **<0.001** | 0.84(0.64-1.10) | 0.210 | 0.61(0.44-0.85) | **0.003** | 0.47(0.33-0.67) | **<0.001** |
| Shockable vs. Non-shockable rhythm | 1.57(1.07-2.31) | **0.022** | 1.21(0.90-1.63) | 0.195 | 1.23(0.84-1.82) | 0.291 | 1.71(1.13-2.60) | **0.012** |

Significance are shown in bold.

**Supplementary Table S2.**

| Outcome | Group | Estimated probability | 95% CI for est.prob. |  |
| --- | --- | --- | --- | --- |
|  |  |  | LB | UB |
| **Pre-hospital ROSC** | No access attempt | 7.54% | 6.51% | 8.71% |
|  | IO w/o E | 12.80% | 10.27% | 15.84% |
|  | IO w/ E | 13.26% | 10.22% | 17.03% |
|  | IV w/o E | 10.56% | 8.69% | 12.77% |
|  | IV w/ E | 10.94% | 8.80% | 13.54% |
|  | Failed IV | 4.60% | 2.63% | 7.92% |
| **Survival over 2 hours** | No access attempt | 21.45% | 19.79% | 23.20% |
|  | IO w/o E | 26.80% | 23.31% | 30.60% |
|  | IO w/ E | 30.48% | 26.09% | 35.27% |
|  | IV w/o E | 22.71% | 20.06% | 25.60% |
|  | IV w/ E | 26.04% | 22.83% | 29.52% |
|  | Failed IV | 18.78% | 14.49% | 23.98% |
| **Survival to discharge** | No access attempt | 9.83% | 8.66% | 11.13% |
|  | IO w/o E | 13.75% | 11.16% | 16.83% |
|  | IO w/ E | 22.18% | 18.02% | 27.00% |
|  | IV w/o E | 7.72% | 6.20% | 9.58% |
|  | IV w/ E | 13.02% | 10.68% | 15.79% |
|  | Failed IV | 8.05% | 5.30% | 12.03% |
| **Good CPC** | No access attempt | 6.82% | 5.84% | 7.94% |
|  | IO w/o E | 7.68% | 5.79% | 10.12% |
|  | IO w/ E | 12.44% | 9.33% | 16.42% |
|  | IV w/o E | 5.61% | 4.31% | 7.28% |
|  | IV w/ E | 9.22% | 7.23% | 11.69% |
|  | Failed IV | 3.45% | 1.80% | 6.49% |

Abbreviation: est. prob., estimate probability

